# Unraveling the spatiotemporal spread of COVID-19 in Brazil through spatial network connectivity

**DOI:** 10.1101/2021.11.16.21266414

**Authors:** Ligia V Barrozo, Christopher Small

## Abstract

**Background:** Describing and understanding the process of diffusion can allow local managers better plan emergence scenarios. Thus, the main aim of this study was to describe and unveil the spatiotemporal patterns of diffusion of the COVID-19 in Brazil from February 2020 until April 2021.

**Methods:** This is a retrospective purely observational ecologic study including all notified cases and deaths. We used satellite-derived night light imagery and spatiotemporal Empirical Orthogonal Function analysis to quantify the spatial network structure of lighted development and the spatiotemporal transmission of the pathogen through the network.

**Results:** The more populous state capitals within the largest network components presented higher frequency of deaths and earlier onset compared to the increasing numbers of smaller, less populous municipalities trending toward lower frequency of deaths and later onset. By week 48 2020, the full network was almost completely affected. Cases and deaths showed a distinct second wave of wider geographic expansion beginning in early November 2020.

**Conclusions:** The spatiotemporal diffusion in Brazil was characterized by an intertwined process of overseas relocation, hierarchical network transmission and contagious effects. A rapid response as the immediate control of all ports, airports and borders combined with mandatory quarantine are critical to retard disease diffusion.

## 1. Introduction

Since the beginning of the COVID-19 pandemic, the disease already killed more than 597,948 people in Brazil until early October 2021. This tragic result can be explained by a series of failures stemming from the belief that collective immunity would be achieved without the need for mass vaccination (1). Leaving aside the reasons that led the country to a late response to the pandemic, comprehending the spatiotemporal pattern of diffusion is of great value since for any unknown infectious agent it takes time for the complexity of the disease to be partially unraveled. In a globalized world, the emergence of new pandemics is unpredictable yet likely. Unveiling the initial diffusion mechanism can allow local managers to take evidence-based strategic preventive action before the entire territory is affected by a new pathogen.

The seminal theory of spatial diffusion set forth by Hägerstrand in 1953 has contributed to understanding the space-time process involved when a disease is disseminated to new locations (2). This process can be classified in the following patterns: relocation, contagious, hierarchical, network and, mixed diffusions (3). However, the spatiotemporal pattern that a same disease may develop varies from context to context even at the same scale. At the national scale, in Italy, for instance, COVID-19 presented a spatial diffusion that can be classified as contagious (4), which is strongly influenced by the friction of distance (e.g., time involved increases with distance) with gradual spread from the north of the country (5,6). In Germany, the spread of COVID-19 did not follow the well-known spatial patterns but from super spreading events and relocation diffusion (7). Some studies on the spread in Brazil have attempted to model the main factors that drove its pace and spatial diffusion (8,9). However, they were based on choropleth maps using large units of geographic analysis, such as states or microregions (10), obscuring the hierarchy structure between cities that can affect spatial connectivity more than distance. Another inherent constraint related to data aggregation in the geographic unit is the modifiable areal unit problem (MAUP) (11), that gives different results depending on the scale of aggregation (12) and changes in the limits of the geographic units. To mitigate these observational constraints, a finer spatial resolution is necessary to represent the network structure which influences the mobility pattern and the real location of population settlements within the administrative units. To resolve these issues, we use night light imagery from the Visible Infrared Imaging Radiometer Suite (VIIRS) Day/Night Band (DNB) sensor on the NASA/NOAA Suomi satellite. In contrast to the spatially varying resolution of Brazil’s 5,570 municipality units, the VIIRS sensor provides uniform 700 m spatial resolution for the entire country. Using this more detailed night light imagery, it is possible to identify the spatial network structure of lighted development as a proxy for the distribution of ambient population to analyze the spatiotemporal evolution of COVID-19 confirmed cases and deaths in Brazil. The network structure is quantified in terms of rank-size distributions of spatially continuous lighted areas, depicting the spatial networks of varying degrees of connectivity within which population are distributed (13). Here we describe the spatiotemporal diffusion of COVID-19 at the municipality scale in Brazil, projecting the aggregated COVID-19 cases and deaths per capita onto the spatial network structure to identify the most important characteristics of the epidemic’s spread. We do not intend to model the mechanism of spread, but to understand its spatiotemporal transmission based on spatially explicit observations. Identifying the main diffusion process can support the definition of the distribution of health units and supplies in the large Brazilian territory for eventual new waves of COVID-19 or another epidemic.

## 2. Materials and methods

This is a retrospective purely observational ecologic study including all notified cases and deaths from COVID-19 from February 25^th^, 2020, to April 16^th^, 2021, by municipality in Brazil.

### Epidemiologic and population data

All epidemiologic data used in this analysis are publicly available and aggregated by geographic unit, municipality of residence not including identifiable individual information, eliminating the need for approval by an ethics committee, as there is no violation of confidentiality.

Daily updates of confirmed cases and deaths from COVID-19 by municipality in Brazil are available at Brazil.IO Project (14). The Brazilian Ministry of Health used different case definitions throughout the pandemic, whose definitions were systematized by Cavalcante et al. (15). Cases data in Brazil are believed to be highly underreported (16). One reason is that in the beginning of the pandemic, the population was oriented to avoid seeking health care in the case of light symptoms. In this sense, only severe cases were reported. Another reason is related to the absence of a mass testing as strategy in the management of the epidemic. However, even with these constraints we analyzed confirmed case and death data because positivity in seroprevalence surveys correlated well geographically with reported mortality due to COVID-19 (17), together they allow us to have a better picture of the diffusion. Population data by municipality for 2020 was estimated by the Brazilian Institute of Geography and Statistics (18) which also provides the digital cartographic database of Brazilian municipalities (19).

To understand the relocation diffusion for the entrance of the virus in the country, we conducted online research on the description of the first case notified by each municipality that reported a case in the first month of the pandemic. This research included epidemiological bulletins from municipalities and/or news from online newspapers.

### VIIRS Night Light

The Visible Infrared Imaging Radiometer Suite (VIIRS) sensor has imaged anthropogenic night light from the NASA/NOAA NPP Suomi satellite since 2012. The digital radiance data are used to produce monthly and annual global composites of temporally stable nighttime lights (20). Cloud-free composites of the VIIRS Day-Night Band (DNB) data provide monthly average brightness (radiance) in units of nW/(cm^2^ sr). Additional thresholding is used to remove ephemeral lights (e.g., fires) and background noise to produce gridded stable lights products (20). The data and documentation are available from: https://payneinstitute.mines.edu/eog-2/viirs/.

The VIIRS stable night light product is gridded at 0.004° (∼500 m @ Equator), oversampling the 700m native resolution of the VIIRS sensor. We reproject the geographic grids to Molleweide equal area grids with a uniform spatial resolution of 500×500 meters. Because radiance of night light varies over several orders of magnitude, we quantify the degree of spatial connectivity by applying low luminance thresholds to the Log_10_ of radiance (nW/(cm^2^ sr)). Higher luminance thresholds attenuate dimmer lights thereby reducing the number and area of network components as well as the spatial connectivity of the network as a whole. The resulting spatial network for the high connectivity threshold of 10^−0.5^ nW/(cm^2^/sr) is shown in comparison the sum of COVID-19 deaths aggregated by municipality, in Figure 1.

**Figure 1:**
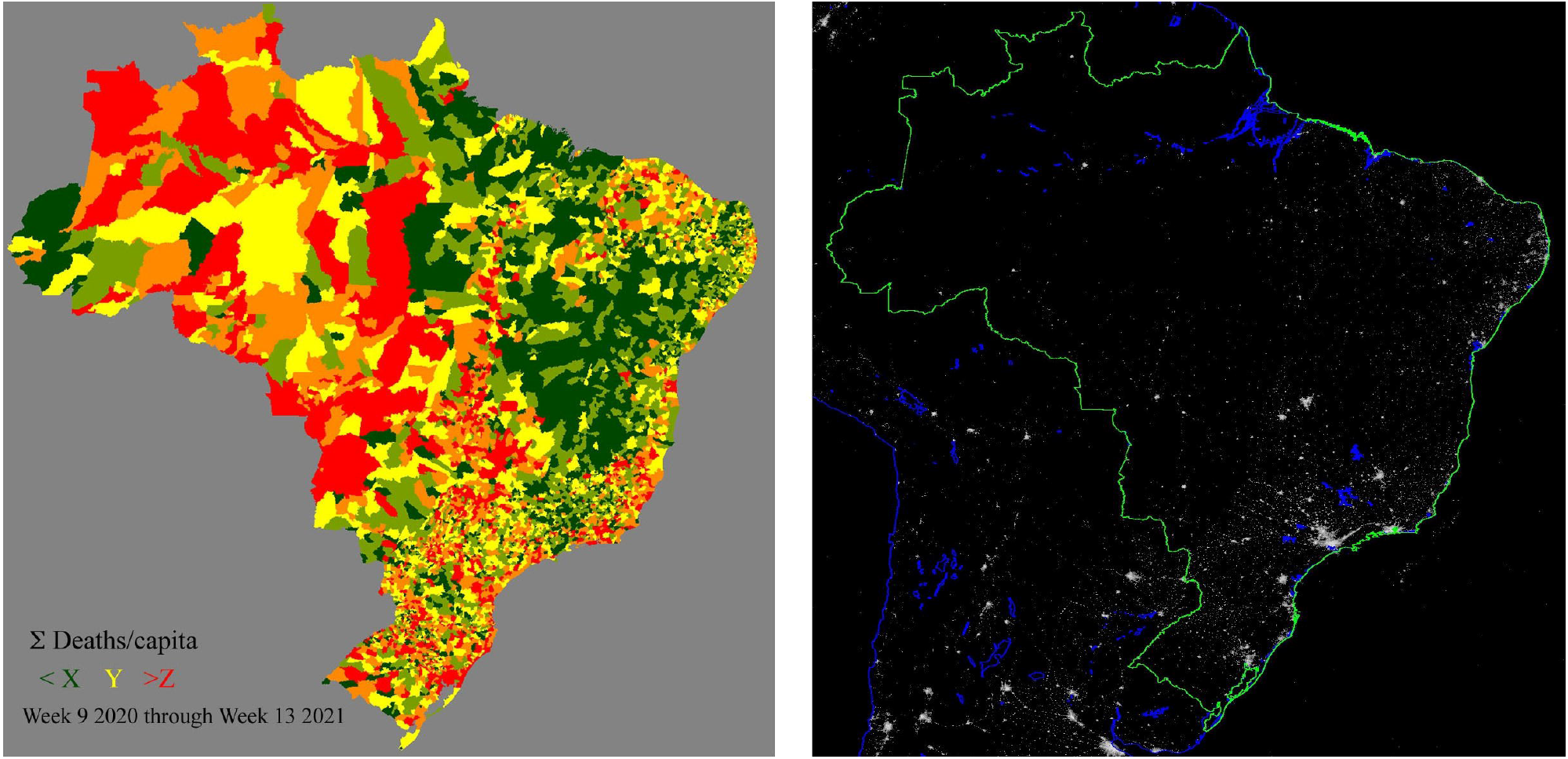
COVID-19 deaths/capita and night light brightness. Brazil’s 5 570 municipalities (left) span 4 orders of magnitude in population and 5 orders of magnitude in area so provide a crude depiction of geographic distribution of the population. Average night light brightness (right) in June 2020 from VIIRS is imaged at uniform ∼700 m spatial resolution so provides a more precise distribution of most of the population at a scale much finer than even the smallest municipality. While the smaller, dimmer lights are not visible at the scale of this image, the 625 000 km2 spatial network of lighted settlements contains ∼15 000 spatially contiguous components spanning 5 orders of magnitude in area.

### Analysis

This spatiotemporal network analysis follows the procedure described in detail by Small et al. (21) and Small and Sousa (13). The methodology combines the spatiotemporal Empirical Orthogonal Function (EOF) analysis described by Small (22) with the multi-threshold spatial network characterization described by Small et al. (23) and refined by Small and Sousa (24). The result of the spatiotemporal EOF analysis is a temporal feature space in which independent dimensions (principal components) represent different temporal patterns that most efficiently capture the diversity of trajectories present in the data. Within this feature space, units with similar temporal trajectories (in this study, daily deaths by municipality) cluster together to provide a concise depiction of the range of temporal trajectories in the data. The low order principal component values for each unit (municipality) can be projected back into geographic space onto the more spatially detailed night light network to represent the spatial distribution of temporal trajectories in their geographic context. The spatial network characterization of the night light data quantifies the size and shape of each spatially contiguous network component, allowing the characteristics of the temporal trajectories to be mapped onto the characteristics of the network structure to provide insight into the relationship between the spatial and temporal dimensions of the evolution of the pandemic within the network. Because COVID-19 deaths are known with greater accuracy than confirmed cases, and not subject to variations in testing, we use the more conservative measure of deaths per capita per day by municipality for the spatiotemporal EOF analysis. However, to illustrate the upper bound scenario for network transmission, we use the less conservative combination of the high spatial connectivity network and the number of confirmed cases per capita per day. The combination of case and death data, and the low, medium and high connectivity of networks allows for 6 different scenarios to be considered to provide both liberal and conservative estimates as needed. For brevity, we present only the most conservative (deaths) and liberal (cases + high connectivity network) scenarios here.

## 3. Results

In the first month of the pandemic in Brazil, from February 26 to March 25, 2020, 221 municipalities reported their first cases from all the 27 Federative Units (FUs) of the country (Supplementary Material 1). Overseas relocation was the process of entrance in 19 FUs (Table 1). Of the 221 municipalities that reported the first case in February/March 2020, 41.18% of the cases were imported, 30.32% were classified as local transmission, 1.8% as community transmission and 26.7% were not disclosed or were unknown. Local transmission was the main entrance in 7 FUs with residents who had visited São Paulo (visitors from Acre, Rondônia, Roraima and, Maranhão), Rio de Janeiro (visitor from Pará), Fortaleza (visitor from Tocantins) and, Belém (visitor from Amapá). On March 20, 2020, the Ministry of Health of Brazil declared community transmission throughout the country.

**Table 1.**
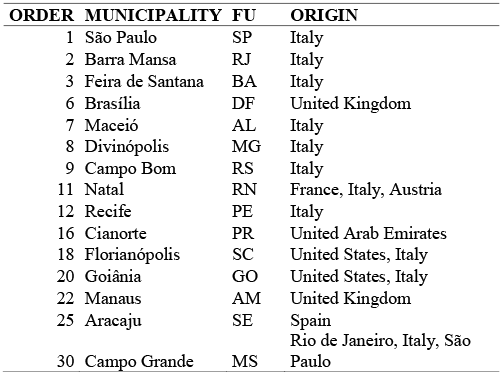
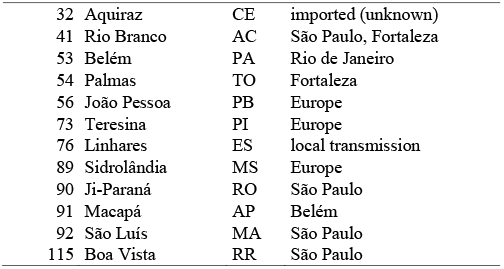
Municipalities of the 27 Brazilian Federative Units that notified the first case of COVID-19, among the 221 municipalities reporting in the first month

The temporal progression of the COVID-19 pandemic in Brazil is summarized in Figure 2. While confirmed cases appear to show four distinct peaks, the latter three are much less distinct in reported deaths. Log_10_ of cases and deaths show similar quasi-linear slopes in the first two months of the pandemic, suggesting comparable rates of exponential growth in the spatial extent of the pathogen. Both clearly show a distinct second wave of geographic expansion beginning in early November 2020. The maximum number of municipalities simultaneously reporting cases and deaths occurred in the epidemiological weeks 10 (3,766 municipalities) and 13 (1,104 municipalities) of 2021, respectively.

**Figure 2:**
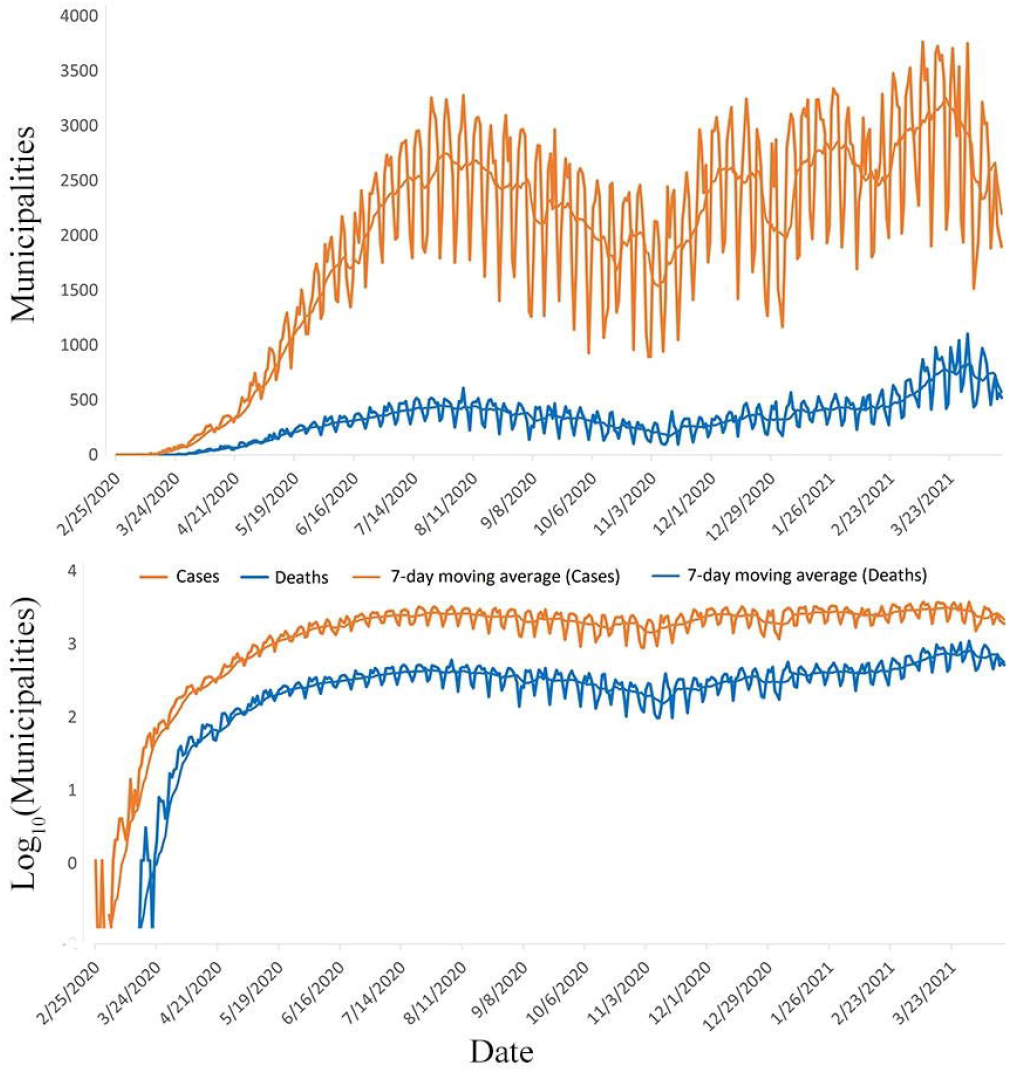
Daily numbers of municipalities reporting confirmed cases and deaths from COVID-19. Confirmed cases suggest four distinct peaks of expansion (top), although the latter three peaks are less distinct in the number of municipalities reporting deaths. Plotting Log10 of the same time series shows comparable exponential increases in cases and deaths in the first two months, asymptotically stabilizing before diminishing till the beginning of a second peak in early November 2020 which reached its maximum in late March 2021. Plotting number of municipalities rather than total number of cases or deaths shows the geographic dispersal more effectively.

The temporal feature space of the first and second principal components of the daily deaths time series illustrates the spatiotemporal evolution of COVID-19 diffusion in Brazil. Figure 3 shows a skewed distribution of PC values with the more populous state capitals corresponding to higher frequency of deaths (+ PC1) and earlier onset (- PC2), in stark contrast to increasing numbers of less populous municipalities trending toward lower frequency of deaths (- PC1) and later onset (+ PC2). Increasing frequency of deaths takes the form of larger numbers of deaths (per 1,000 residents) per day, and more continuous sequences of days reporting deaths – in contrast to smaller, less populous municipalities reporting lower numbers of deaths occurring more intermittently between days without reported deaths. Earlier onset takes the form of greater numbers of deaths increasing more rapidly earlier in 2020, while later onset takes the form of both delayed occurrence of first deaths as well as later increases of larger numbers of deaths in 2021.

**Figure 3:**
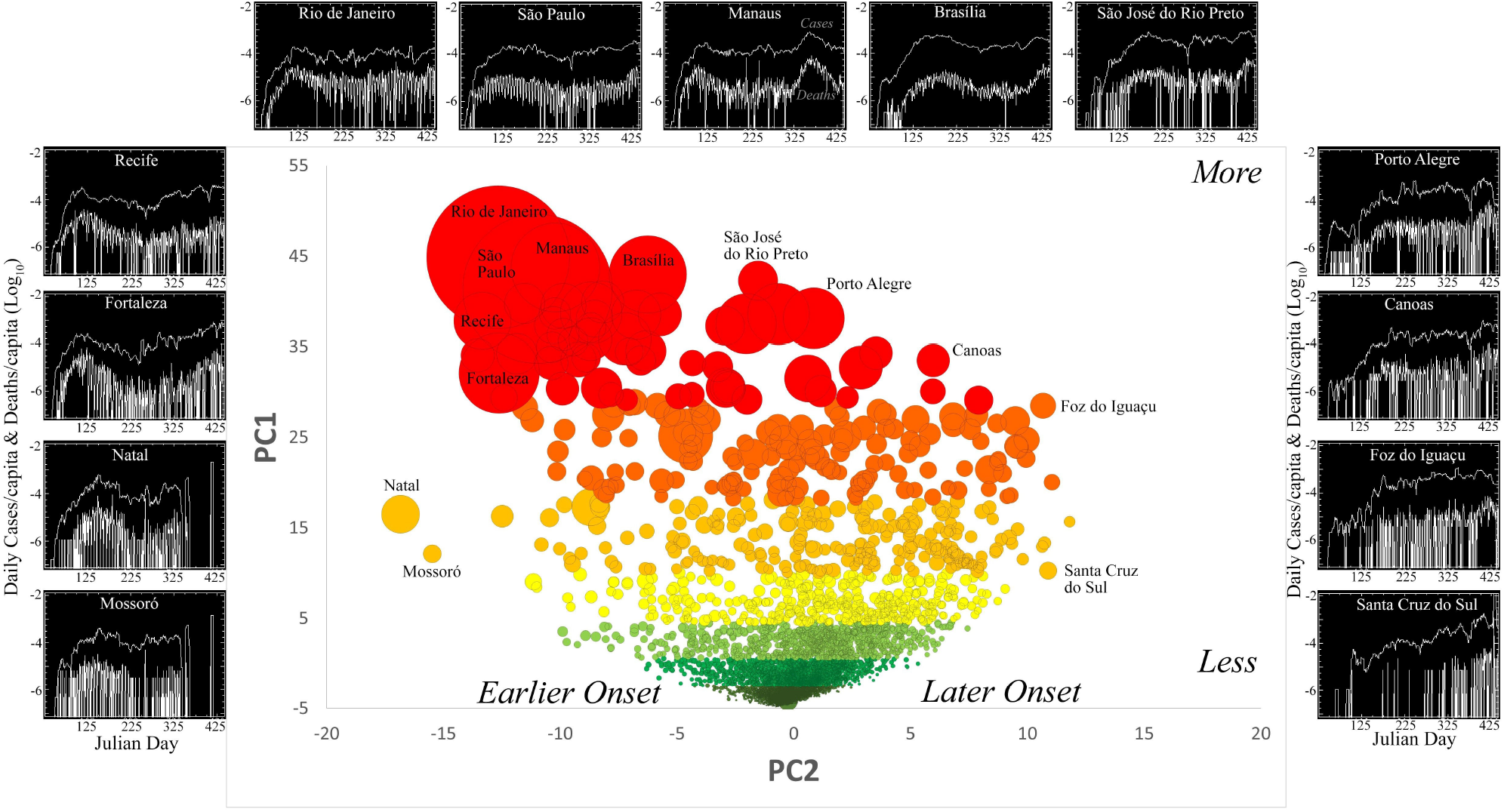
Temporal feature space for deaths per capita trajectories. The first and second Principal Components account for 18% and 2% of total variance (respectively), suggesting a wide variety of trajectories. Symbol size is proportional to the total deaths per municipality. The larger, earlier onsets and higher frequencies of deaths while smaller are skewed toward latter onsets. Deaths shown as 7 day moving average.

When temporal frequency of deaths (PC1) by municipality is projected onto the spatial network of lighted development the effect of the network structure on the spatiotemporal evolution of transmission is clear. Figure 4 shows the spatial network of night light for Brazil’s Southeast Corridor at full spatial resolution. The color of individual network components corresponds to the PC1 value of the municipality within which they occur (for smaller components contained within a larger municipality) or the distribution of PC1 values of the municipalities within components spanning multiple municipalities. It is immediately apparent that the highest temporal frequencies of deaths (red) occur in the few largest network components (e.g., São Paulo, Rio de Janeiro) while the lowest temporal frequencies of deaths (green) occur in the many smaller, more isolated, components.

**Figure 4:**
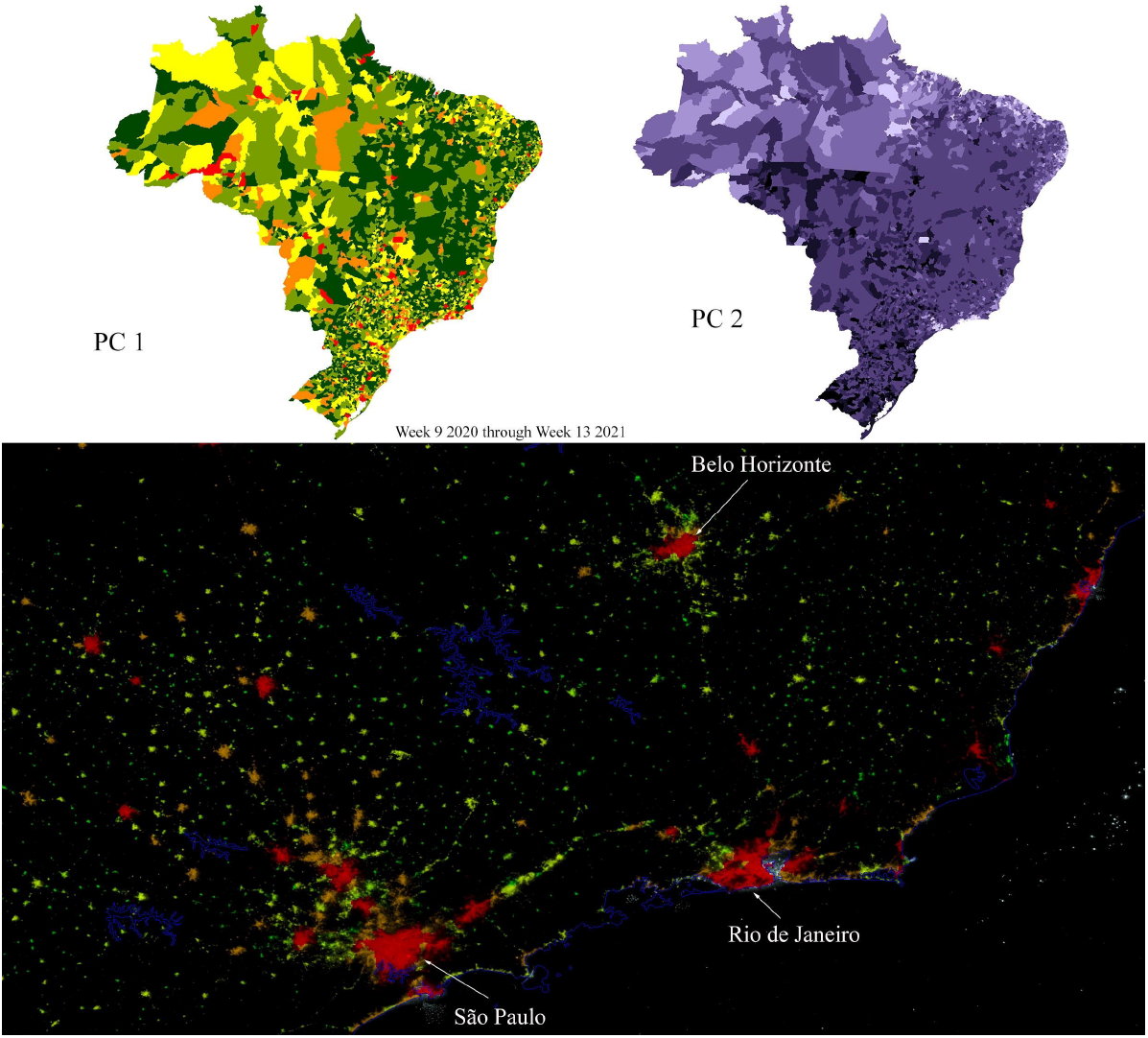
Spatiotemporal Principal Components show frequency (PC1 top left) and onset timing (PC2 top right) of the COVID-19 death trajectories shown in Figure 3. When PC values at the municipality scale are projected onto the spatial network of night light the trajectories can be mapped in much greater detail. The network+trajectory map of PC1, shown at full resolution for the Southeast Corridor, clearly shows higher frequency of deaths/capita in the larger components.

The spatiotemporal transmission of the SARS-CoV2 pathogen through the spatial network of lighted settlements (and population) can be quantified as the temporal progression of network components first reporting confirmed cases. If the spatial network structure is segmented as a binary complement of infected and uninfected municipalities on each day, the temporal progression of number and size of infected components can be tracked through time as transmission through the network occurs. This spatiotemporal network transmission dynamic can be illustrated with an animation of daily confirmed cases for the early stage of the pandemic (http: https://youtu.be/YEOrkKms6X0). This spatiotemporal network transmission can be summarized concisely using time series of rank-size distributions of infected network components. Rank-size distributions quantify the spatial scaling of settlement networks as continua ranging from small numbers of large network components down to large numbers of small network components. Figure 5 shows the temporal evolution of number of infected components and total area of infected network, as well as the progression of rank-size distributions of infected network components. The former clearly shows the exponential progression of both number and total area of infected network components. The latter shows the parallel increase in infected component area and the downward progression of this increase from fewer larger components to more smaller components as the infected network grows through time. By week 48, the full network is almost completely affected. However, the remaining uninfected municipalities occur at the peripheries of the few largest network components (upper tail) as well as in large numbers of the smallest components (lower tail).

**Figure 5:**
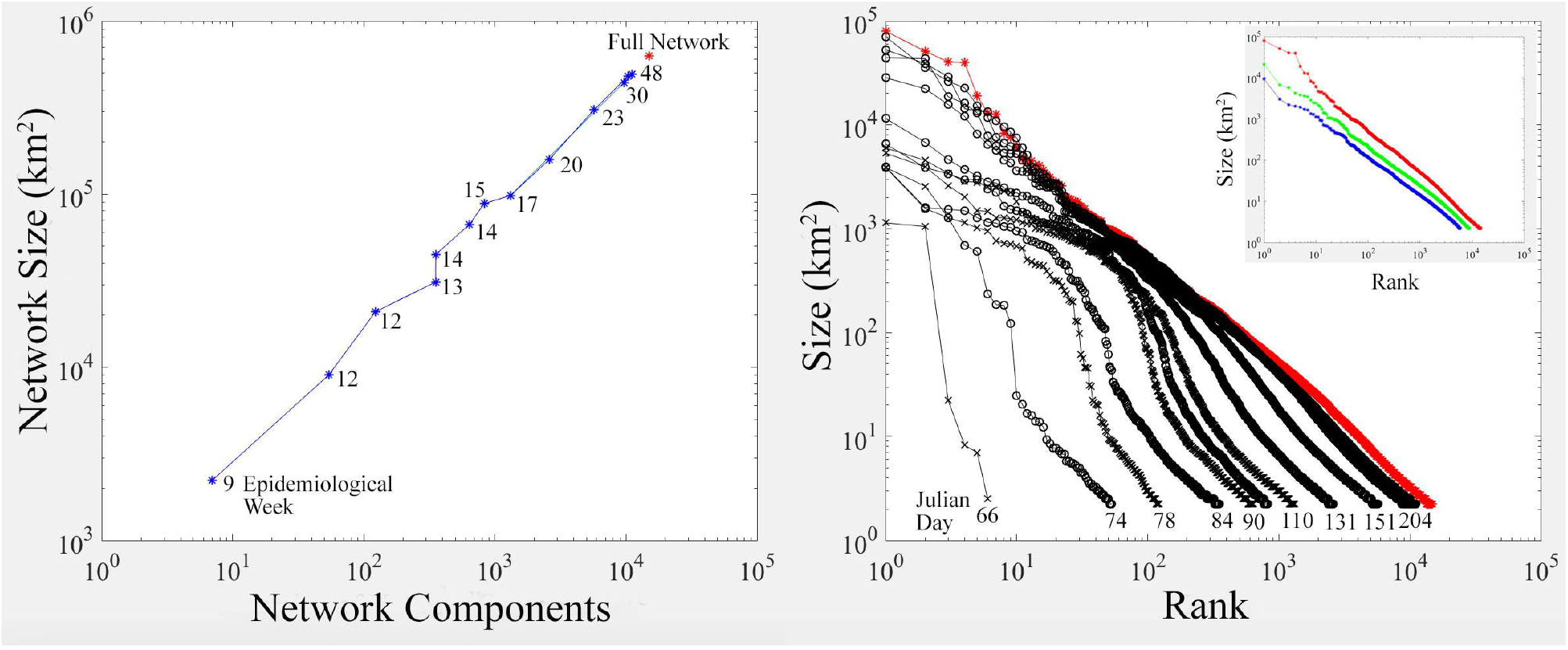
Exponential growth of the spatial network of confirmed cases (infection+detection). The number of components and total area of the network (left) increase exponentially throughout 2020. As the pathogen spread from the largest cities within the most interconnected components, outward to exponentially increasing numbers of smaller components (right), the shoulder of the rank-size distribution propagates downward. By March 2021 almost the entire network has been exposed to the pathogen. The full network (red) corresponds to the higher connectivity case with the lowest luminance threshold, so this represents the least conservative depiction of propagation efficiency. The inset shows the rank-size distributions of the low, medium and high connectivity networks. All have similar scaling with slopes near -1.

## 4. Discussion

This population-based study on the spatiotemporal spread of COVID-19 cases and mortality in Brazil shows that the disease simultaneously entered in the country in 19 of 27 FUs. The entire process of diffusion in the country took nine months with the widest geographic expansion occurring in the epidemiological weeks 10 and 13 2021, for cases and deaths, respectively. Both the most and less developed regions, Southeast and North Regions, have been reached at the same time. However, Manaus, capital of the State of Amazonas in the North Region, was far more hard hit due to the lack of oxygen cylinders in hospitals (25) and the collapse of the funeral system.

The COVID-19 spatiotemporal diffusion in Brazil was characterized by an intertwined process of overseas relocation, hierarchical network, and contagion effects. Here we call attention to the fact that except for seven FUs (mainly from the Northern Region), imported cases were the main form of introduction of the virus in the first 30 days, in 38.9% of the municipalities that reported their first cases. Not only São Paulo and Rio de Janeiro received the virus via imported cases but 70% of the FUs. After entering the country, the diffusion follows a leapfrog spread across network components in which the distance was not a barrier. On the contrary, spread by contagion (in neighboring municipalities) is noticed only after more distant cities have reported their first cases or deaths. The Gould’s (4) image of “stones” being dropped into the pond of the territory, each one producing a wave-like pulse of contagion to municipalities in its area of influence can be applied to the process of COVID-19 diffusion in Brazil. Coarser scales of aggregation to the municipality level would not allow to identify this spatial pattern because the locations of the population within the municipality is not known. Projecting municipality temporal trajectories onto more detailed night light imagery avoids the MAUP in this cartographic scale because it represents the actual location and spatial connectivity of lighted settlements in which the vast majority of the population resides.

Although the entire diffusion took long time to be completed, the most vulnerable Region had its first case on March 13 2020 (26), only 16 days after the first notified case of the country in São Paulo. Amazonas is particularly vulnerable to COVID-19 due to the large presence of indigenous who are known to be a risk group (26) and due to the large percentage of dwellers in precarious settlements (27). Immediate restrictions on the movement of people by air, road and river could have delayed the rapid breakthrough of the disease and, perhaps, avoided the two collapses in this Region.

Some limitations of this study deserve attention. As all studies based on secondary data, our analysis may be biased due to the delay in reporting the first case or death of the municipality, which can affect the temporal dimension of the network analysis. However, if the delay occurred more in the smaller municipalities, the order of the diffusion has likely not been affected.

## 5. Conclusions

This observational study provides new spatiotemporal constraints to prior simulations of transmission in Brazil. Here we intend to quantify the diffusion process of the disease in a large territory because the understanding of this mechanism can provide support for subside strategic plans for the control of health emergencies caused by infectious diseases of prompt transmission. SARS-CoV-2 entered in Brazil simultaneously through different airports of which it was dissipated following road network, cities hierarchy and neighboring metropolitan cities at the same time. A rapid response as the immediate control of all ports, airports and borders combined with mandatory quarantine as of January 30, 2020, when cases had already been reported in at least 21 countries (28) could have reduced the number of victims of the disease in Brazil.

## Data Availability

All data produced in the present study are available upon reasonable request to the authors.

https://youtu.be/YEOrkKms6X0

## CRediT author statement

L.V.B. and C.S. developed the conceptualization and worked on data curation. C.S. conducted the EOF and network analysis and L.V.B. conducted online research. L.V.B. wrote the first draft of the paper and C.S. contributed to subsequent revisions and approved the final version.

## Appendix A. Supplementary data

The following are the Supplementary data to this article: Supplementary Material 1. Order of the first notified cases of COVID-19 in Brazil by municipality in the first month, Federative Unit, origin, type of transmission and source of information.

